# Subsequent waves of viral pandemics, a hint for the future course of the SARS-CoV-2 pandemic

**DOI:** 10.1101/2020.07.10.20150698

**Authors:** Fabian Standl, Karl-Heinz Jöckel, Bernd Kowall, Börge Schmidt, Andreas Stang

## Abstract

**Background:** It is unknown if the SARS-CoV-2 pandemic will have a second wave. We analysed published data of five influenza pandemics (such as the Spanish Flu and the Swine Flu) and the SARS-CoV-1 pandemic to describe whether there were subsequent waves and how they differed.

**Methods:** We reanalysed literature and WHO reports on SARS-CoV-1 and literature on five influenza pandemics. We report frequencies of second and third waves, wave heights, wavelengths and time between subsequent waves. From this, we estimated peak-to-peak ratios to compare the wave heights, and wave-length-to-wave-length ratios to compare the wavelengths differences in days. Furthermore, we analysed the seasonality of the wave peaks and the time between the peak values of two waves.

**Results:** Second waves, the Spanish Flu excluded, were usually about the same height and length as first waves and were observed in 93% of the 57 described epidemic events of influenza pandemics and in 42% of the 19 epidemic events of the SARS-CoV-1 pandemic. Third waves occurred in 54% of the 28 influenza and in 11% of the 19 SARS-CoV-1 epidemic events. Third waves, the Spanish Flu excluded, usually peaked higher than second waves with a peak-to-peak ratio of 0.5.

**Conclusion:** While influenza epidemics are usually accompanied by 2nd waves, this is only the case in the minority of SARS-Cov1 epidemics.

## Introduction

It is currently unknown whether, when and to what extent the SARS-CoV-2 pandemic will be followed by subsequent waves. It was stated that the last blueprint like pandemics were in 1957 (Asian Flu) and 1968 (Hong Kong Flu).(1) It was furthermore speculated, by referring to the Spanish Flu (1918-1920), that a second wave of the current SARS pandemic could be far worse.(2) We revisited influenza pandemics of the last 130 years and the SARS-CoV-1 pandemic to find out, how often subsequent waves were observed, if subsequent waves peaked higher and how many days lay between the peaks of the waves.

## Material and Methods

A pandemic consists of one or more “epidemic events” at different places. We analysed available data on corona virus and influenza pandemics and found one corona and five influenza pandemics. In total, since 1889 there were seven influenza pandemics.(3) We did not find accessible data on the waves of the 1900 pandemic and the Russian Flu of 1977 and therefore did not incorporate it in this work. In total, we collected data for 57 epidemic events from the literature for the influenza pandemics and 19 for SARS-CoV-1: three epidemic events for the Russian Flu (RF) from 1889 to 1895 (H3N8),(4) 29 events for the Spanish Flu (SF) from 1918 to 1920 (H1N1),(4-13) four for the Asian Flu (AF) from 1957 to 1960 (H2N2),(14-16) ten for the Hong Kong Flu (HKF) (H3N2) from 1968 to 1970,(17, 18) 19 for the SARS-CoV-1 pandemic (COV1) from 2002 to 2004(19) and 11 for the Swine Flu (SWF) from 2009 to 2010 (H1N1).(3, 20-25) As SF represented 29 of 57 influenza epidemic events, we ran two further analysis: one for SF alone and one for the remaining influenza epidemic events. For COV1, we collected 11 events from the literature and eight from the WHO reports.(26-31)

Relevant literature was identified by PubMed. The search terms we used were: “pandemic wave”, “pandemic waves”, “Russian Flu waves”, “Spanish Flu Waves”, “Asian Flu waves”, “Hong Kong Flu waves”, “Swine Flu waves”, “SARS pandemic”, “SARS pandemic waves”. In addition, we manually searched for further publications from the reference lists of identified papers. For inclusion into our study, publications had to display time series graphs of the epidemic waves. Only for the WHO reports on COV1 we made an exception, as we regarded all of them to be relevant.

Reports on the severity of influenza epidemics methodologically varied. For example, some reports mentioned the total number of deaths per day, other reports reported the number of infected people per 100.000 population or the number of “influenza like illnesses” (ILI) – some papers displayed one or more methods for the same epidemic event.(32) We collected data on the occurrence of subsequent waves, the peak heights of waves, the wavelength and the time between peaks. From this, we estimated the time between peaks, the ratios of wavelengths and the ratios of peaks by dividing the peak of one wave and the peak of a subsequent wave (peak-to-peak ratio and corresponding interquartile range [IQR]); the same approach was applied for the wave-length-to-wave-length ratio. This means we divided the respective numbers of a first wave, stated in a paper, by the numbers of a second wave and the numbers of a second wave by the numbers of a third wave. Subsequent waves were identified by visual inspection of the epidemic curves. Peaks that could clearly be separated from each other were regarded as two subsequent waves. We estimated the wavelength by visually analysing the epidemic curves. As images varied again from paper to paper, we used several approaches for identifying the wavelength. In general, we regarded the wavelength to be the time, the epidemic curve needed, after leaving a normal level, to reach a minimum with a turning point or a minimum with normal values thereafter. For the calculation in days, we defined each month to have 30 days and each year to have 365 days. Above this, we calculated the time between the wave peaks (Δt) and used midpoints of time intervals when papers did not provide daily information but e.g. weekly or monthly data. Although seven out of 57 epidemic events contained a fourth wave, we only included the first three waves in our analysis.(4, 13) Finally, we analysed the seasonality by assigning the corresponding season (spring, summer, fall, winter) to the peaks of the epidemic events.(33)

For COV1, we additionally analysed all of the consecutive WHO-reports (17^th^ March 2003 to 11^th^ July 2003) with regard to the cumulative number of cases. We re-calculated daily new cases by subtracting subsequent cumulative numbers of cases. Although most of the reports were published on a daily basis, few reports covered two or more days. We therefore calculated the new cases per day by dividing the number of new cases per report by the number of days which passed between one report and its subsequent report. Sometimes the numbers of the cumulative cases have been revised down, which resulted in negative values for new cases per day. We have distributed the negatively coded case numbers proportionally to the daily cases until that date. However, especially for China, some excessively high values remained. It is likely that those values reflect cumulative cases rather than daily new cases. After we reviewed all 39 plots produced for the respective countries and areas in the reports, we identified eight charts that display epidemic events (with a local transmission and not only sporadic, imported cases) and therefore fulfil the scope of this work. To dampen the random fluctuation of case numbers in the eight plots, we used a seven-day moving average for smoothing.

## Results

Overall, 53 (93%) out of 57 influenza epidemic events showed a second wave and 34 out of 57 (60%) showed a third wave. 42% of the first waves peaked in summer, 49% of the second waves in fall and 59% of the third waves peaked in winter (Table 1). First and third waves were about the same length with a median duration of 90 [56, 140] respectively 100 [73, 165] days, whereas the second waves had a median duration of 120 days [91, 175]. The time between the first and the second peak (136 days [118, 273]) was shorter than the time between the second and the third peak (305 days [133, 440]). Usually the second waves peaked higher than the other two waves (peak-to-peak ratio of 0.9 [0.4, 1.4] respectively 1.2 [0.5, 2.5]) (Table 2).

**Table 1:**
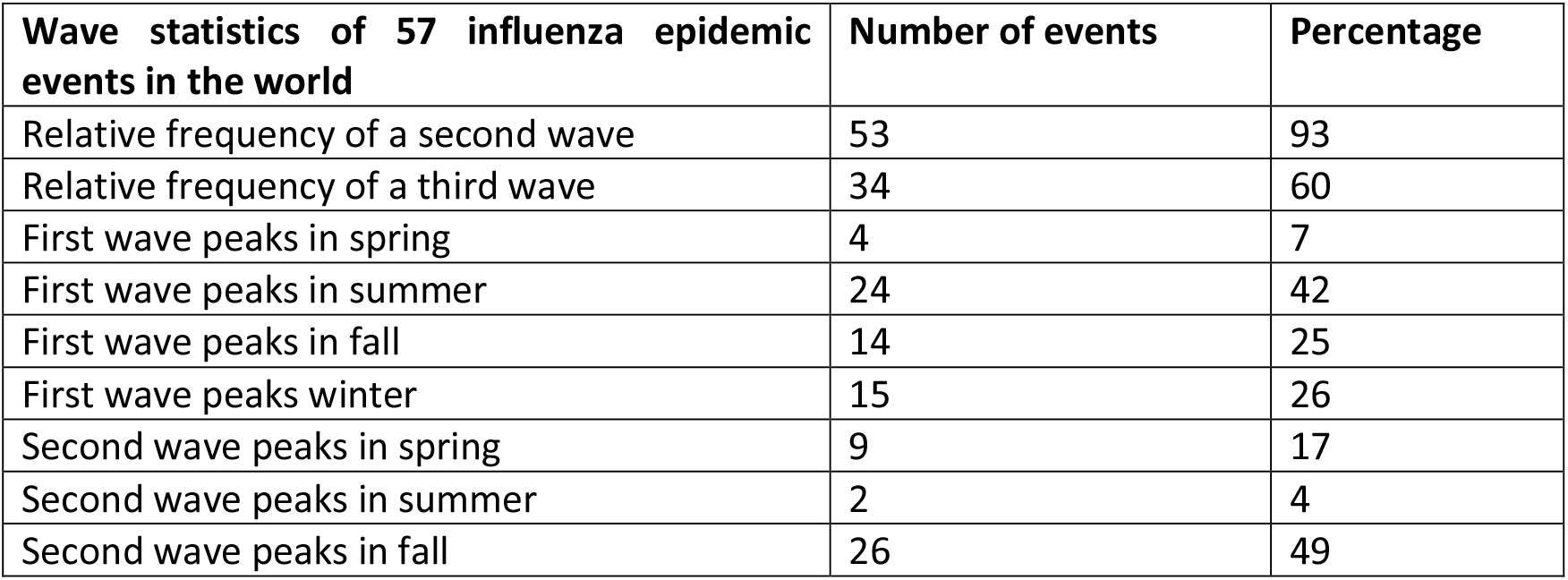
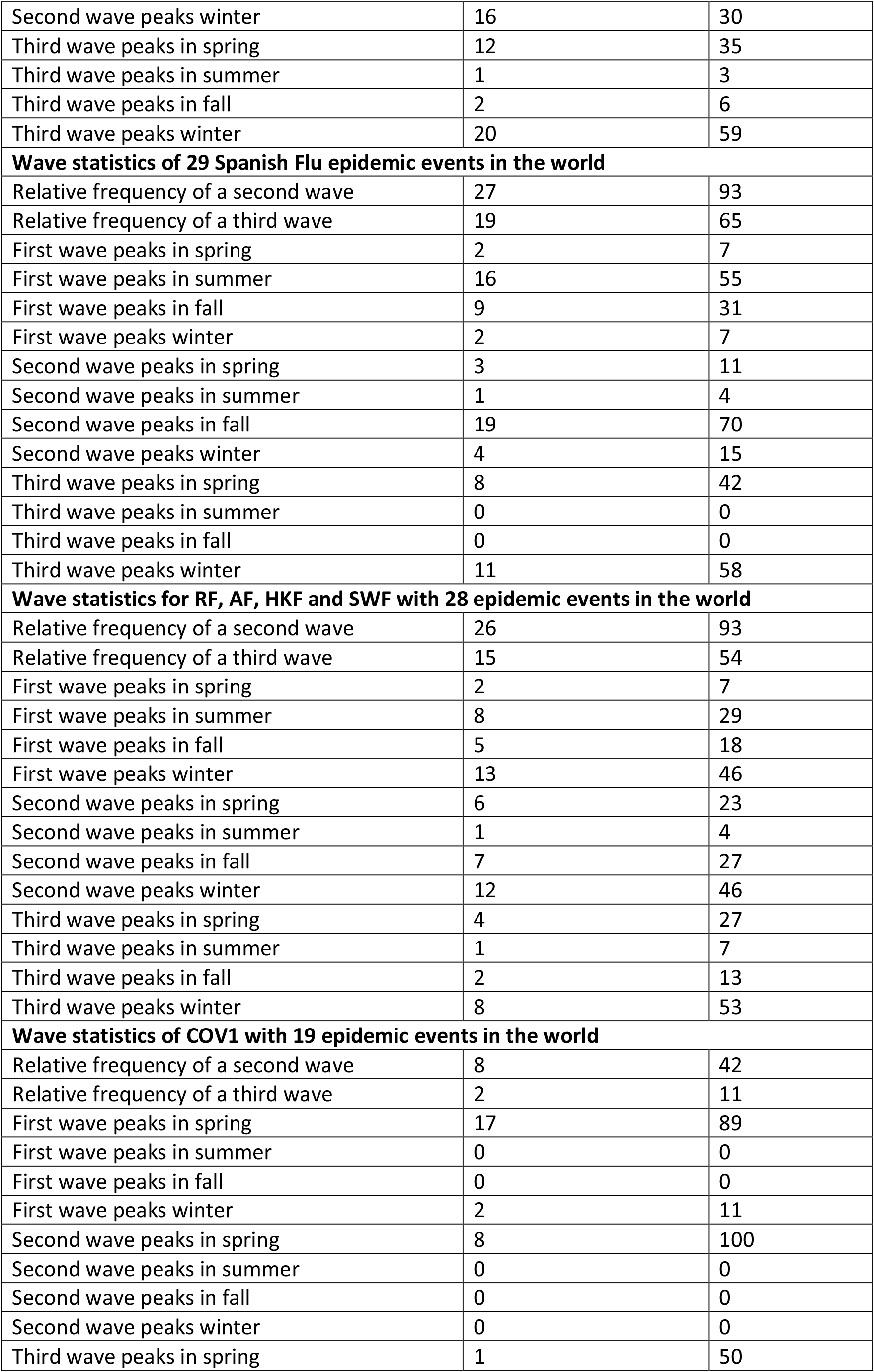
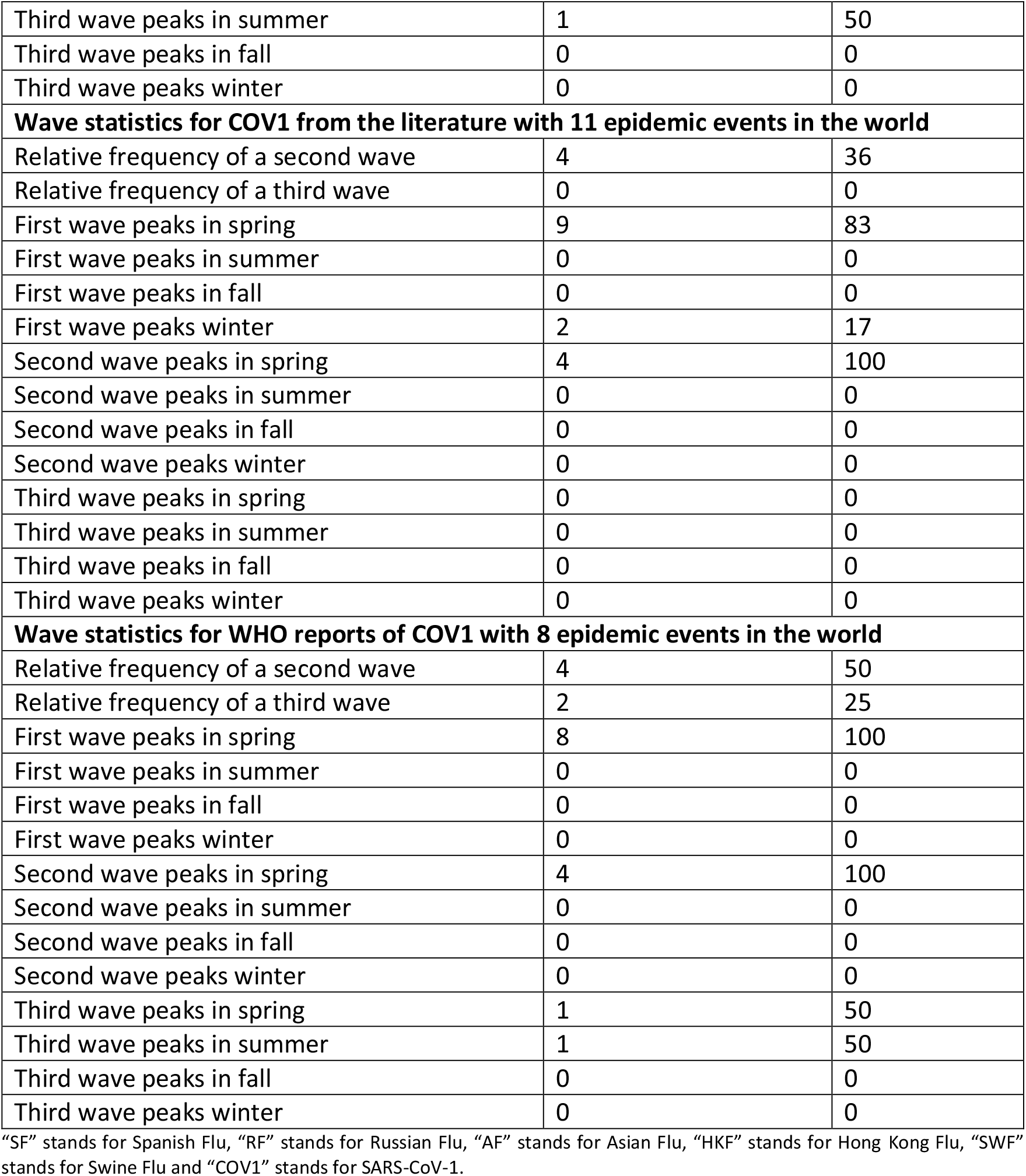
Relative frequency and peak season.

| **Wave statistics of 57 influenza epidemic events in the world** | **Number of events** | **Percentage** |
| --- | --- | --- |
| Relative frequency of a second wave | 53 | 93 |
| Relative frequency of a third wave | 34 | 60 |
| First wave peaks in spring | 4 | 7 |
| First wave peaks in summer | 24 | 42 |
| First wave peaks in fall | 14 | 25 |
| First wave peaks winter | 15 | 26 |
| Second wave peaks in spring | 9 | 17 |
| Second wave peaks in summer | 2 | 4 |
| Second wave peaks in fall | 26 | 49 |
| Second wave peaks winter | 16 | 30 |
| Third wave peaks in spring | 12 | 35 |
| Third wave peaks in summer | 1 | 3 |
| Third wave peaks in fall | 2 | 6 |
| Third wave peaks winter | 20 | 59 |
| **Wave statistics of 29 Spanish Flu epidemic events in the world** |  |  |
| Relative frequency of a second wave | 27 | 93 |
| Relative frequency of a third wave | 19 | 65 |
| First wave peaks in spring | 2 | 7 |
| First wave peaks in summer | 16 | 55 |
| First wave peaks in fall | 9 | 31 |
| First wave peaks winter | 2 | 7 |
| Second wave peaks in spring | 3 | 11 |
| Second wave peaks in summer | 1 | 4 |
| Second wave peaks in fall | 19 | 70 |
| Second wave peaks winter | 4 | 15 |
| Third wave peaks in spring | 8 | 42 |
| Third wave peaks in summer | 0 | 0 |
| Third wave peaks in fall | 0 | 0 |
| Third wave peaks winter | 11 | 58 |
| **Wave statistics for RF, AF, HKF and SWF with 28 epidemic events in the world** |  |  |
| Relative frequency of a second wave | 26 | 93 |
| Relative frequency of a third wave | 15 | 54 |
| First wave peaks in spring | 2 | 7 |
| First wave peaks in summer | 8 | 29 |
| First wave peaks in fall | 5 | 18 |
| First wave peaks winter | 13 | 46 |
| Second wave peaks in spring | 6 | 23 |
| Second wave peaks in summer | 1 | 4 |
| Second wave peaks in fall | 7 | 27 |
| Second wave peaks winter | 12 | 46 |
| Third wave peaks in spring | 4 | 27 |
| Third wave peaks in summer | 1 | 7 |
| Third wave peaks in fall | 2 | 13 |
| Third wave peaks winter | 8 | 53 |
| **Wave statistics of COV1 with 19 epidemic events in the world** |  |  |
| Relative frequency of a second wave | 8 | 42 |
| Relative frequency of a third wave | 2 | 11 |
| First wave peaks in spring | 17 | 89 |
| First wave peaks in summer | 0 | 0 |
| First wave peaks in fall | 0 | 0 |
| First wave peaks winter | 2 | 11 |
| Second wave peaks in spring | 8 | 100 |
| Second wave peaks in summer | 0 | 0 |
| Second wave peaks in fall | 0 | 0 |
| Second wave peaks winter | 0 | 0 |
| Third wave peaks in spring | 1 | 50 |
| Third wave peaks in summer | 1 | 50 |
| Third wave peaks in fall | 0 | 0 |
| Third wave peaks winter | 0 | 0 |
| **Wave statistics for COV1 from the literature with 11 epidemic events in the world** |  |  |
| Relative frequency of a second wave | 4 | 36 |
| Relative frequency of a third wave | 0 | 0 |
| First wave peaks in spring | 9 | 83 |
| First wave peaks in summer | 0 | 0 |
| First wave peaks in fall | 0 | 0 |
| First wave peaks winter | 2 | 17 |
| Second wave peaks in spring | 4 | 100 |
| Second wave peaks in summer | 0 | 0 |
| Second wave peaks in fall | 0 | 0 |
| Second wave peaks winter | 0 | 0 |
| Third wave peaks in spring | 0 | 0 |
| Third wave peaks in summer | 0 | 0 |
| Third wave peaks in fall | 0 | 0 |
| Third wave peaks winter | 0 | 0 |
| **Wave statistics for WHO reports of COV1 with 8 epidemic events in the world** |  |  |
| Relative frequency of a second wave | 4 | 50 |
| Relative frequency of a third wave | 2 | 25 |
| First wave peaks in spring | 8 | 100 |
| First wave peaks in summer | 0 | 0 |
| First wave peaks in fall | 0 | 0 |
| First wave peaks winter | 0 | 0 |
| Second wave peaks in spring | 4 | 100 |
| Second wave peaks in summer | 0 | 0 |
| Second wave peaks in fall | 0 | 0 |
| Second wave peaks winter | 0 | 0 |
| Third wave peaks in spring | 1 | 50 |
| Third wave peaks in summer | 1 | 50 |
| Third wave peaks in fall | 0 | 0 |
| Third wave peaks winter | 0 | 0 |

**Table 2:** Wave statistics results in the world.

| **Wave statistics of 57 influenza epidemic events in the world** | n | Median | IQR | Min. Max. |
| --- | --- | --- | --- | --- |
| Length of first wave in days | 57 | 90 | 56, 140 | 21, 330 |
| Days between the peak of the first and the peak of the second wave | 53 | 136 | 118, 273 | 36, 761 |
| Peak height of first wave divided by peak height of second wave | 53 | 0.9 | 0.4, 1.4 | 0.1, 12.0 |
| Length of second wave in days | 53 | 120 | 91, 175 | 14, 280 |
| Wave length of first wave divided by wave length of second wave | 53 | 0.8 | 0.6, 1.0 | 0.2, 4.8 |
| Days between the peak of the second and the peak of the third wave | 34 | 305 | 133, 440 | 49, 517 |
| Peak height of second wave divided by peak of third wave | 34 | 1.2 | 0.5, 2.5 | 0.2, 11.0 |
| Length of third wave in days | 34 | 100 | 73, 165 | 45, 330 |
| Wave length of second wave divided by wave length of third wave | 34 | 1.0 | 0.8, 1.5 | 0.2, 2.5 |
| **Wave statistics of 29 Spanish Flu epidemic events in the world** |  |  |  |  |
| Length of first wave in days | 29 | 90 | 61, 120 | 45, 330 |
| Days between the peak of the first and the peak of the second wave | 27 | 136 | 122, 183 | 74, 488 |
| Peak height of first wave divided by peak height of second wave | 27 | 0.6 | 0.3, 1.1 | 0.1, 5.9 |
| Length of second wave in days | 27 | 120 | 90, 180 | 60, 240 |
| Wave length of first wave divided by wave length of second wave | 27 | 0.7 | 0.5, 1.0 | 0.3, 3.8 |
| Days between the peak of the second and the peak of the third wave | 19 | 335 | 123, 488 | 92, 517 |
| Peak height of second wave divided by peak of third wave | 19 | 2.3 | 1.3, 3.8 | 0.5, 11.1 |
| Length of third wave in days | 19 | 90 | 61, 150 | 45, 195 |
| Wave length of second wave divided by wave length of third wave | 19 | 1.3 | 0.9, 1.6 | 0.4, 2.5 |
| **Wave statistics for RF, AF, HKF and SWF with 28 epidemic events in the world** |  |  |  |  |
| Length of first wave in days | 28 | 105 | 67, 148 | 21, 266 |
| Days between the peak of the first and the peak of the second wave | 26 | 161 | 105, 365 | 36, 761 |
| Peak height of first wave divided by peak height of second wave | 26 | 1.1 | 0.7, 1.8 | 0.2, 12.0 |
| Length of second wave in days | 26 | 116 | 102, 150 | 14, 280 |
| Wave length of first wave divided by wave length of second wave | 26 | 0.87 | 0.6, 1.1 | 0.2, 4.8 |
| Days between the peak of the second and the peak of the third wave | 15 | 304 | 203, 420 | 49, 431 |
| Peak height of second wave divided by peak of third wave | 15 | 0.5 | 0.3, 1.0 | 0.2, 3.3 |
| Length of third wave in days | 15 | 122 | 75, 180 | 63, 330 |
| Wave length of second wave divided by wave length of third wave | 15 | 0.8 | 0.6, 1.0 | 0.2, 1.8 |
| **Wave statistics of COV1 with 19 epidemic events in the world** |  |  |  |  |
| Length of first wave in days | 19 | 50 | 23, 68 | 10, 150 |
| Days between the peak of the first and the peak of the second wave | 8 | 49 | 19, 70 | 14, 72 |
| Peak height of first wave divided by peak height of second wave | 8 | 1.0 | 0.5, 1.2 | 0.3, 1.6 |
| Length of second wave in days | 8 | 36 | 27, 45 | 13, 63 |
| Wave length of first wave divided by wave length of second wave | 8 | 1.1 | 0.5, 1.3 | 0.2, 1.7 |
| Days between the peak of the second and the peak of the third wave | 2 | - | - | 28, 43 |
| Peak height of second wave divided by peak of third wave | 2 | - | - | 0.6, 1.5 |
| Length of third wave in days | 2 | - | - | 10, 37 |
| Wave length of second wave divided by wave length of third wave | 2 | - | - | 0.5, 1.3 |
| **Wave statistics for COV1 from the literature with 11 epidemic events in the world** |  |  |  |  |
| Length of first wave in days | 11 | 88 | 45, 88 | 25, 150 |
| Days between the peak of the first and the peak of the second wave | 4 | 70 | 64, 72 | 58, 72 |
| Peak height of first wave divided by peak height of second wave | 4 | 1.1 | 0.7, 1.4 | 0.3, 1.6 |
| Length of second wave in days | 4 | 39 | 36, 52 | 35, 63 |
| Wave length of first wave divided by wave length of second wave | 4 | 1.3 | 0.9, 1.5 | 0.5, 1.7 |
| Days between the peak of the second and the peak of the third wave | - | - | - | - |
| Peak height of second wave divided by peak of third wave | - | - | - | - |
| Length of third wave in days | - | - | - | - |
| Wave length of second wave divided by wave length of third wave | - | - | - | - |
| **Wave statistics for WHO reports of COV1 with 8 epidemic events in the world** |  |  |  |  |
| Length of first wave in days | 8 | 22 | 16, 65 | 10, 75 |
| Days between the peak of the first and the peak of the second wave | 4 | 19 | 16, 30 | 14, 39 |
| Peak height of first wave divided by peak height of second wave | 4 | 0.8 | 0.5, 1.0 | 0.5, 1.1 |
| Length of second wave in days | 4 | 27 | 17, 41 | 13, 49 |
| Wave length of first wave divided by wave length of second wave | 4 | 0.8 | 0.4, 1.1 | 0.2, 1.2 |
| Days between the peak of the second and the peak of the third wave | 2 | - | - | 28, 43 |
| Peak height of second wave divided by peak of third wave | 2 | - | - | 0.6, 1.5 |
| Length of third wave in days | 2 | - | - | 10, 37 |
| Wave length of second wave divided by wave length of third wave | 2 | - | - | 0.5, 1.3 |

27 (93%) out of 29 SF epidemic events showed a second wave and 19 out of 29 (65%) showed a third wave. 55% of the first waves peaked in summer, 70% of the second waves in fall and 58% of the third waves peaked in winter or, for 42%, in spring (Table 1). First and third waves were about the same length with a median duration of 90 [61, 120] respectively 90 [61, 150] days, whereas the second waves had a median duration of 120 days [90, 180]. The median time between the first and the second peak (136 days [122, 183]) was shorter than the time between the second and the third peak (355 days [123, 488]). Usually the second waves peaked higher than the other two waves (peak-to-peak ratio of 0.6 [0.3, 1.1] respectively 2.3 [1.3, 3.8]) (Table 2). Beside this SF showed a clear multimodality for the delta time from peak of the second wave to the peak of the third wave with 92, 457 and 488 days. Another clear multimodality was observed for the length of the third wave with 61, 90 and 150 days (Figure 1).

**Figure 1:**
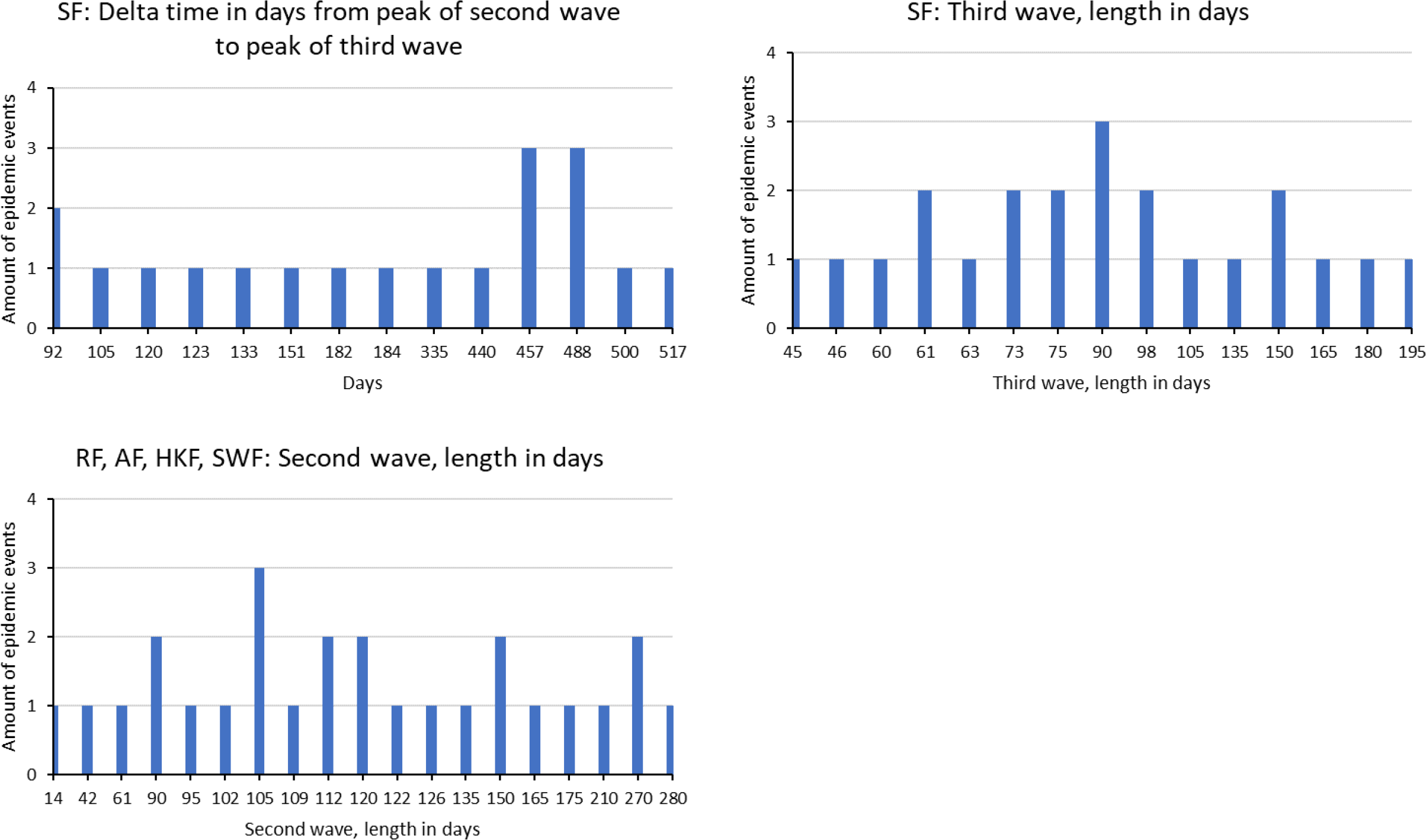
Multimodalities in influenza pandemics. “SF” stands for Spanish Flu, “RF” stands for Russian Flu, “AF” stands for Asian Flu, “HKF” stands for Hong Kong Flu and “SWF” stands for Swine Flu.

Combined for RF, AF, HKF and SWF in 26 (93%) out of 28 epidemic events a second wave could be observed and a third wave in 15 out of 28 (54%) epidemic events. 46% of the first waves peaked in winter and so did 46% of the second waves and 53% of the third waves (Table 1). First waves had a median duration of 105 [67, 148] days, second waves usually took a little longer with 116 days [102, 150] and third waves tended to be a little longer than second waves with 122 days [75, 180]. The time between the first and the second peak (161 days [105, 365]) was shorter than the time between the second and the third peak (304 days [203, 420]). Usually the peaks of the first and the second waves were about the same height (peak-to-peak ratio of 1.1 [0.7, 1.8]), whereas the peaks of the third waves were smaller 0.5 [0.3, 1.0] (Table 2). Beside this, a multimodality was observed for the length of the second wave in days with 90, 105, 112, 120, 150 and 270 days (Figure 1).

Overall, COV1 showed a second wave in 8 (42%) out of 19 epidemic events and a third wave in 2 out of 19 (10%) epidemic events. 89% of the first waves peaked in spring as well as 100% of the second waves. The two observed third waves peaked in spring and summer (Table 1). First waves were longer (50 days [23, 68]) than second waves (36 days [27, 45]) and the two third waves were 10 and 37 days long. The time between the first and the second peak was 49 days [19, 70]. The first and the second wave peaked at similar heights (peak-to-peak ratio of 1.0 [0.5, 1.3]) (Table 2).

COV1 literature showed a second wave in 4 (35%) out of 11 epidemic events and no third waves. 83% of the first waves peaked in spring as well as 100% of the second waves (Table 1). First waves were longer (88 days [45, 88]) than second waves (39 days [36, 52]). The time between the first and the second peak was 70 days [64, 72]. The first and the second wave peaked at similar heights (peak-to-peak ratio of 1.1 [0.7, 1.4]) (Table 2).

The analysis of the WHO-COV1 reports showed a second wave in 4 (50%) out of 8 epidemic events and a third wave in 2 out of 8 (25%) epidemic events. 100% of the first waves peaked in spring as well as 100% of the second waves. The two observed third waves peaked in spring and summer (Table 1). First waves were shorter (22 days [16, 65]) than second waves (27 days [17, 41]) and the two third waves were 10 and 37 days long. The time between the first and the second peak was 19 days [16, 30]. The first and the second wave peaked at similar heights (peak-to-peak ratio of 0.8 [0.5, 1.0]) (Table 2).

By plotting the results of the WHO reports, we found that in Canada a total of 249 cases were observed. The main epidemic happening stretched over about 90 days. During this time, three waves occurred with peaks on the 1^st^ of April 2003, the 22^nd^ of April 2003 and the 4^th^ of June 2003. It was uncertain whether Canada had two or three waves (Figure 2a), as the peak of the 22^nd^ of April might still belong to the first wave.

**Figure 2a:**
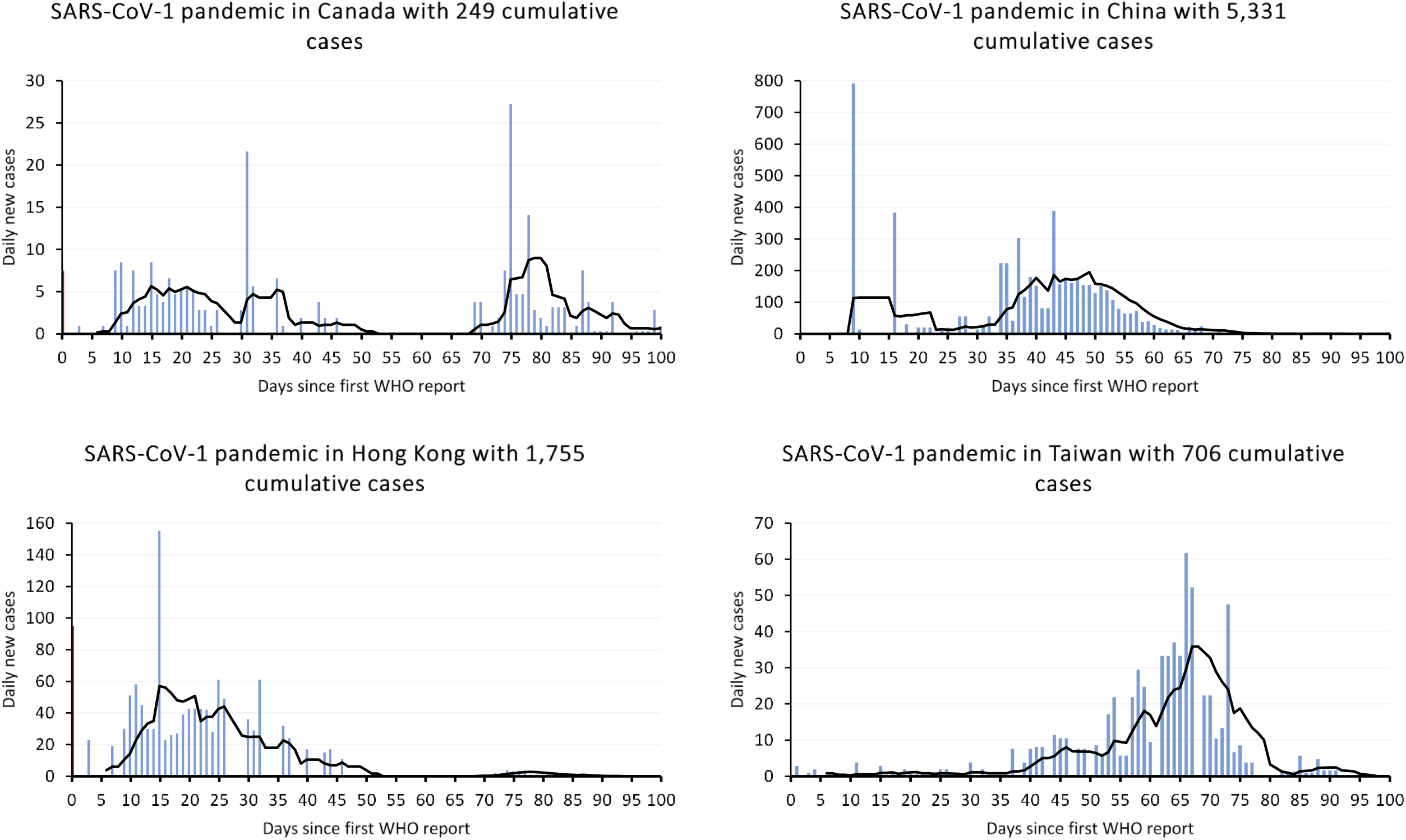
SARS-CoV-1 pandemic in Canada, China, Hong Kong and Taiwan. The blue columns show the daily new cases. The black line is the seven-day simple moving average. The red column at the very beginning are likely to be cumulative cases from an earlier point in time. The zero value on the X-Axis marks the day of the onset of the WHO reports, which is the 17^th^ of March 2003.

**Figure 2b:**
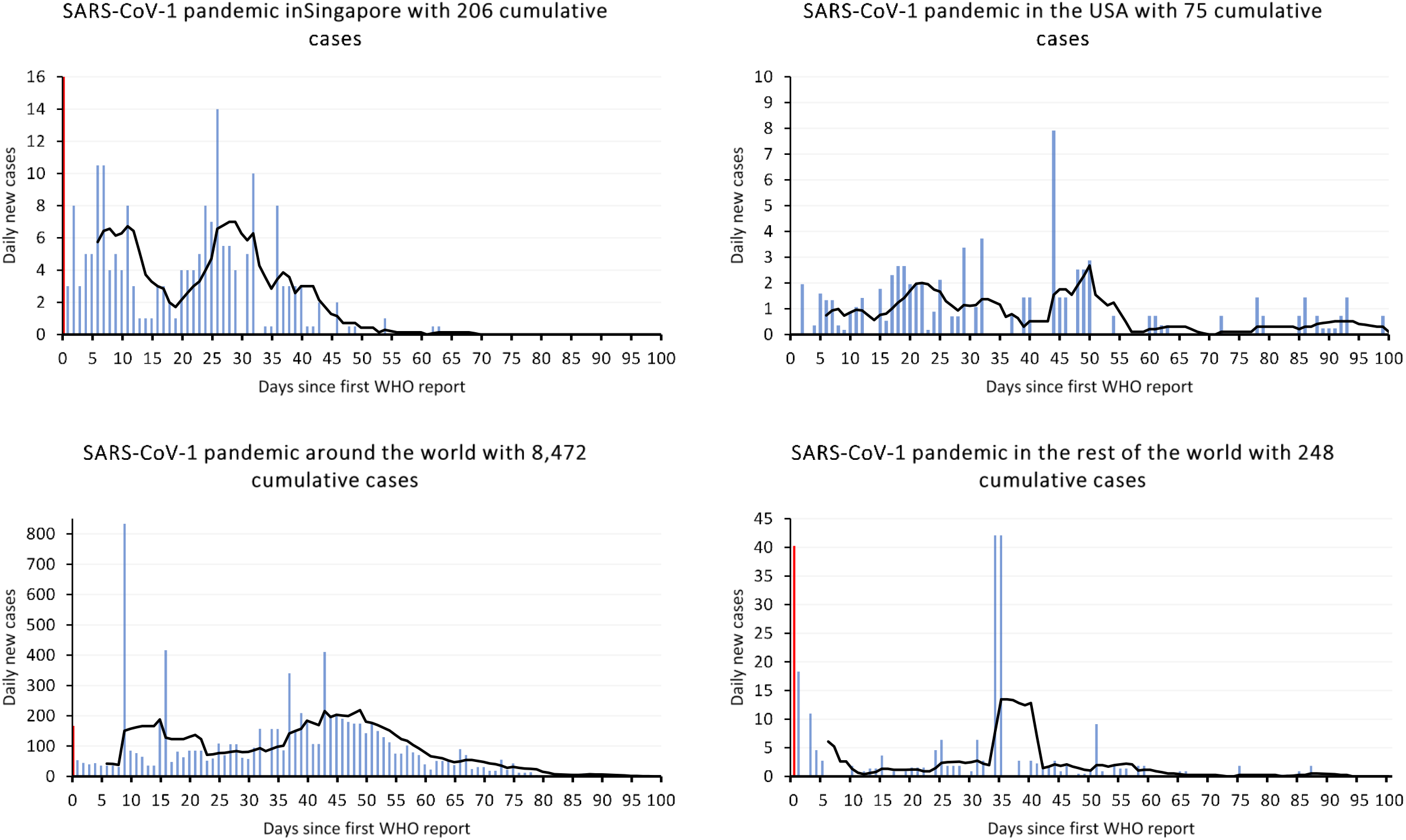
SARS-CoV-1 in Singapore, USA globally and the rest of the world. The blue columns show the daily new cases. The black line is the seven-day simple moving average. The red column at the very beginning are likely to be cumulative cases from an earlier point in time. The zero value on the X-Axis marks the day of the onset of the WHO reports, which is the 17^th^ of March 2003. “Rest of the world” represents the epidemic happening that was observed after having subtracted the happenings in Canada, China, Hong Kong, Taiwan, Singapore and USA from the global, cumulative happening.

For China we found a total of 5,331 cases, which stretched – according to the reporting – over about 60 days. The outlying cases counts on day 9 and 16 were most likely an artefact due to the accumulation of cases registered the previous days. If one ignores the outlying days 9 and 16, China suffered from one wave that peaked on day 49 of the epidemic (May 5, 2003) and another peak of an earlier wave may have peaked on day ten of the epidemic (March 27, 2003) (Figure 2a).

For Hong Kong a total number of 1,755 cases was observed which are distributed over the epidemic wave displayed in Figure 2a. The wave peaked on the 1^st^ of April 2003 and stretched over about 45 days.

For Taiwan we found a total of 706 cases which were distributed over a single wave, that stretched over about 45 days and peaked on 23^rd^ of May 2003. We found no further hints for a subsequent epidemic happening, apart from the small wave that stretched from about day 85 to day 95 (Figure 2a).

For Singapore we found a total of 206 cases which clearly stretched over 2 waves. The epidemic happening stretched over at least 50 days. It is not deducible from the WHO data whether there was an epidemic happening before day zero. However, if there was another wave before day zero, this must have been before the 1^st^ of November of 2002, as the WHO reports covered cases from this date on which were reported from day zero onwards. As there was not a sole excessive column at the beginning of the time series, it can be assumed that there was no excessive number reported before day zero. Therefore, it is very likely, that the epidemic happening of COV1 in Singapore started short before day zero. The two peaks of the epidemic happenings were be observed for the 28^th^ of March 2003 and the 14^th^ of April 2003 (Figure 2b).

For the United States of America (USA) we found a total of 75 cases. The main epidemic happening stretched over about 55 days. After the three observable waves, that peaked on the 23^rd^ of March 2003, the 8^th^ of April 2003 and the 6^th^ of May 2003, one could observe further, scattered cases over a timespan of about 45 days. This pattern reminds of an oscillation (Figure 2b).

Globally we found a total of 8,472 cases that stretched over an observable, epidemic happening of about 80 days and which peaked on the 5^th^ of May 2003 (Figure 2b).

After having subtracted the cases of Canada, China, Hong Kong, Taiwan, Singapore and the USA from the global cases, one can calculate the pandemic happening in the rest of the world, which was a total of 248 cases. Figure 2b shows the pandemic footprint in the rest of the world. It could be observed, that it is likely that an epidemic happening might have taken place before day zero. However, from the WHO reports, only one wave was observed that peaked on the 21^st^ of April 2003.

## Discussion

We found for the influenza pandemics and their epidemic footprints around the globe that a second wave occurred in most cases and could usually be observed within a year after the first wave reached its peak. The peaks of a first and a second epidemic wave were about the same height. A third epidemic wave appeared in about 50% of the epidemic events and peaked about one year after the second wave reached its peak. Third waves, the SF excluded, had usually higher peak values than the second waves. Subsequent waves tended to take about 10% longer in days than previous waves.

For COV1 and its epidemic footprints around the globe we found, that a second wave occurred in about half of the cases where an epidemic happening was observed. Usually, a second wave peaked within 70 days after the peak of the first wave. Second waves were about the same height as the first waves, with a downward trend. A third epidemic wave was a rare occurrence and peaked about 35 days after the peak of the second wave. An aggregated view on third waves is currently not possible, however it needs to be mentioned, that third waves were observed. With regard to the wavelength, the calculated ratios show, that first and second waves were about the same length in days.

Combining the results, subsequent waves were observed in small pandemics like COV1, with total cases of about 8,096 globally (official count) and in severe pandemics like SF with a speculation of 100 million deaths globally.(34, 35) Still, it is interesting, that subsequent waves can be observed even in small epidemic happenings. The low Δt between the waves of COV1 might be a result of the relative low peak values, compared to other pandemics displayed above.

We have revisited five influenza-pandemics and the COV1 pandemic with their epidemiological footprints in different parts of the world. Although pandemics are a prominent topic, we found relatively rare accessible data which allow to analyse pandemics in the way we did. Therefore, it must be considered that our sample size is relatively small and that the respective pandemics occurred in a time span of about 120 years. Future works, similar to ours, should improve the comparability of the waves by e.g. calculating the area under the curve of each wave. This approach will make the impact of each wave better assessable and therefore results will be more reliable. As we did not have sufficient data on all pandemics that allowed a homogenous application of this approach, we had to focus on the peaks and the wavelengths. On the other side, this study is the most extensive work on this topic and can provide essential information to improve risk assessments or models. By using the clear and evident methodology future works can easily be compared to this paper.

Generally, epidemiology should reconsider the definition of the term “pandemic” as the only epidemiological part of the WHO definition is, that an illness can be observed in at least two parts of the world.(36) Ioannidis lately stated a similar thought that makes the question deducible whether COV1 with a total of about 8,000 to 8,500 cases globally can really be covered by the same terminology as SF which may have killed 1/18 of the world population.(37, 38)

With regard to the implications and interpretations of our study, the following is deducible: it seems likely that the current SARS-CoV-2 pandemic will have a subsequent wave, which will likely be comparable to the first wave in the sense of the peak-to-peak-ratio. It can be assumed that a second wave may follow the seasonal patterns of influenza pandemics and become epidemic again in fall or winter or, if SARS-CoV-2 follows the patterns of COV1, a second wave might be independent of a seasonal pattern and occur earlier than fall or winter.

## Conclusion

We found that a second and even a third wave usually happened in influenza pandemics. Furthermore, subsequent waves were observed during the COV1 pandemic, but less frequently. Combining both results, subsequent waves for the SARS-CoV-2 pandemic seem to be likely. With regard to the severity it can be assumed that successive waves are likely to be similar to each other with regard to the peak-to-peak-ratios. Furthermore, wavelengths are comparable long and seasonality usually plays an important role. This means that a subsequent wave might peak within one year after the first wave peaked. Reviewing our results, it may be speculated, for SARS-CoV-2, that a subsequent wave may occur in fall respectively winter or earlier. The lesson learned from previous pandemics is, subsequent waves occur, usually peak in winter and are about the same height and length as the previous wave.

## Data Availability

All data are publicly available and can be accessed by using the references.

## Literature

1. “Es wird schlimm werden” [press release]. Internet: Sueddeutsche Zeitung, 28.02.2020 2020.

2. Drosten warnt vor zweiter Corona-Welle [press release]. n-tv.de: ntv, 20.04.2020 2020.

3. Amato Gauci A, Zucs P, Snacken R, Ciancio B, Lopez V, Broberg E, et al. The 2009 A(H1N1) pandemic in Europe. Stockholm: ECDC; 2010.

4. Zürcher K, Zwahlen M, Ballif M, Rieder HL, Egger M, Fenner L. Influenza Pandemics and Tuberculosis Mortality in 1889 and 1918: Analysis of Historical Data from Switzerland. PLoS One. 2016;11(10):e0162575.

5. Morens DM, Taubenberger JK, Harvey HA, Memoli MJ. The 1918 influenza pandemic: lessons for 2009 and the future. Critical care medicine. 2010;38(4 Suppl):e10–20.

6. Palmer CT, Sattenspiel L, Cassidy C. Boats, Trains, and Immunity: The Spread of the Spanish Flu on the Island of Newfoundland. Newfoundland and Labrador Studies. 2007;22(2).

7. VIRAL WAVES Devastating second wave of coronavirus ‘inevitable’ and could hit during flu season [press release]. Internet: The U.S. Sun, 21.04.2020 2020.

8. Curson P, McCracken K. An Australian perspective of the 1918-1919 influenza pandemic. New South Wales public health bulletin. 2006;17(7-8):103–7.

9. How Spanish flu epidemic devastated Wales in 1918 [press release]. Internet: BBC.com, 12.10.2018 2018.

10. Chowell G, Viboud C, Simonsen L, Miller MA, Acuna-Soto R. Mortality patterns associated with the 1918 influenza pandemic in Mexico: evidence for a spring herald wave and lack of preexisting immunity in older populations. J Infect Dis. 2010;202(4):567–75.

11. Olson DR, Simonsen L, Edelson PJ, Morse SS. Epidemiological evidence of an early wave of the 1918 influenza pandemic in New York City. 2005;102(31):11059–63.

12. Chowell G, Erkoreka A, Viboud C, Echeverri-Dávila B. Spatial-temporal excess mortality patterns of the 1918–1919 influenza pandemic in Spain. BMC Infectious Diseases. 2014;14(1):371.

13. Ansart S, Pelat C, Boelle P-Y, Carrat F, Flahault A, Valleron A-J. Mortality burden of the 1918–1919 influenza pandemic in Europe. 2009;3(3):99–106.

14. Cobos AJ, Nelson CG, Jehn M, Viboud C, Chowell G. Mortality and transmissibility patterns of the 1957 influenza pandemic in Maricopa County, Arizona. BMC Infectious Diseases. 2016;16(1):405.

15. Chowell G, Simonsen L, Fuentes R, Flores J, Miller MA, Viboud C. Severe mortality impact of the 1957 influenza pandemic in Chile. 2017;11(3):230–9.

16. Dauer CC. Mortality in the 1957-58 influenza epidemic. Public Health Rep. 1958;73(9):803–10.

17. Jester BJ, Uyeki TM, Jernigan DB. Fifty Years of Influenza A(H3N2) Following the Pandemic of 1968. 2020;110(5):669–76.

18. Viboud C, Grais RF, Lafont BAP, Miller MA, Simonsen L. Multinational Impact of the 1968 Hong Kong Influenza Pandemic: Evidence for a Smoldering Pandemic. J Infect Dis. 2005;192(2):233–48.

19. Bondy SJ, Russell ML, Laflèche JM, Rea E. Quantifying the impact of community quarantine on SARS transmission in Ontario: estimation of secondary case count difference and number needed to quarantine. BMC public health. 2009;9:488.

20. Chakrarat Pittayawonganon HC, Sopon Iamsirithaworn, Pilaipan Puthavathana, Sukhum Chaleysub, Prasert Auewarakul, Somkid Kongyu, Kumnuan Ungchusak, Pasakorn Akarasewi. Monitoring the influenza pandemic of 2009 in Thailand by a community-based survey. Journal of Public Health and Epidemiology 2011;3(4):187–93.

21. Sridhar S, Begom S, Bermingham A, Hoschler K, Adamson W, Carman W, et al. Incidence of Influenza A(H1N1) pdm09 Infection, United Kingdom, 2009–2011. Emerging Infectious Diseases. 2013;19(11):1866 –9.

22. Mummert A, Weiss H, Long L-P, Amigó JM, Wan X-F. A Perspective on Multiple Waves of Influenza Pandemics. PLOS ONE. 2013;8(4):e60343.

23. Centers for Disease Control and Prevention. Severe Illness from 2009 Pandemic Influenza A (H1N1) --- Utah, 2009--10 Influenza Season. Internet: Centers for Disease Control and Prevention (CDC); 2011 30.11.2011. Contract No.: 38.

24. Homaira N, Luby S, Alamgir A, Islam K, Paul R, Abedin J, et al. Influenza-associated mortality in 2009 in four sentinel sites in Bangladesh. Bull World Health Organ. 2012;90.

25. Archer B, Cohen C, Naidoo D, Thomas J, Makunga C, Blumberg L, et al. Interim report on pandemic H1N1 influenza virus infections in South Africa, April to October 2009: Epidemiology and factors associated with fatal cases.. Euro Surveill. 2009;14(42).

26. Xu C, Wang J, Wang L, Cao C. Spatial pattern of severe acute respiratory syndrome in-out flow in 2003 in Mainland China. BMC infectious diseases. 2014;14:3843.

27. Cao W, Fang L, Xiao D. What we have learnt from the SARS epdemics in mainland China? Global Health Journal. 2019;3(3):55–9.

28. Centers for Disease Control and Prevention. Update: Severe Acute Respiratory Syndrome ---Toronto, Canada, 2003. Internet: Centers for Disease Control and Prevention (CDC); 2003 12.06.2003.

29. Svoboda T, Henry B, Shulman L, Kennedy E, Rea E, Ng W, et al. Public Health Measures to Control the Spread of the Severe Acute Respiratory Syndrome during the Outbreak in Toronto. 2004;350(23):2352–61.

30. National Advisory Committee on SARS and Public Health. Chapter 2: Learning from SARS: Renewal of public health in Canada – SARS in Canada: anatomy of an outbreak. Public Health Agency of Canada; 2004.

31. World Health Organization. Cumulative Number of Reported Probable Cases of Severe Acute Respiratory Syndrome (SARS) Internet [Available from: https://www.who.int/csr/sars/country/en/.

32. Pittayawonganon C, Chootrakool H, Iamsirithaworn S, Puthavathana P, Chaleysub S, Auewarakul P, et al. Monitoring the influenza pandemic of 2009 in Thailand by a community-based survey. Journal of Public Health and Epidemiology. 2011;3:187–93.

33. Bikos K, Kher A. Seasons: Meteorological and Astronomical [cited 2020 12.06.]. Available from: https://www.timeanddate.com/calendar/aboutseasons.html.

34. World Health Organization. Summary of probable SARS cases with onset of illness from 1 November 2002 to 31 July 2003. WHO; 2004 21.04.2004.

35. Taubenberger J, Morens D. 1918 Influenza: the Mother of All Pandemics. Emerg Infect Dis. 2006;12(1):15–22.

36. World Health Organization. 4. The WHO pandemic phases. Pandemic Influenza Preparedness and Response: A WHO Guidance Document. Geneva 2009.

37. Johnston R. Historical World Population Data Internet 2015 [updated 21.02.2015. Available from: http://www.johnstonsarchive.net/other/worldpop.html.

38. Bernert J. 250 Expertenstimmen zur Corona-Krise Internet: KenFM; 2020 [Available from: https://kenfm.de/250-expertenstimmen-zur-corona-krise/.

